# Impact of Subthalamic Nucleus Stimulation on Urinary Dysfunction and Constipation in Parkinson’s Disease

**DOI:** 10.1101/2022.05.30.22275772

**Authors:** Asra Askari, Brandon Zhu, Jordan Lam, Kara Wyant, Kelvin Chou, Parag Patil

## Abstract

**Introduction:** The effect of subthalamic nucleus deep brain stimulation (STN DBS) on urinary dysfunction and constipation in Parkinson’s disease (PD) is variable. This study aims to identify potential surgical and non-surgical variables predicting their outcome.

**Methods:** We used the Movement Disorder Society-Unified PD Rating Scale (MDS-UPDRS) Part I to assess urinary dysfunction (item 10) and constipation (item 11) preoperatively and 6-12 months postoperatively. A multiple linear regression model was used to investigate the impact of Global Cerebral Atrophy (GCA) and active electrode contact location on the urinary dysfunction and constipation follow-up score, controlling for age, disease duration, baseline score, motor improvement, and levodopa-equivalent dose changes. An electric field model was applied to localize the maximal effect-site for constipation and urinary dysfunction compared to motor improvement.

**Result:** Among 74 patients, 23 improved, 28 deteriorated, and 23 remained unchanged for urinary dysfunction; 25 improved, 15 deteriorated, and 34 remained unchanged for constipation. GCA score and age significantly predicted urinary dysfunction follow-up score (R^2^ = 0.36, P<0.001). Increasing GCA and age independently were associated with worsening urinary symptoms. Disease duration, baseline constipation score, and anterior active electrode contacts in both hemispheres were significant predictors for constipation follow-up score (R^2^ =0.31, P<0.001). Higher baseline constipation score and disease duration were associated with worsening constipation; anterior active contact location was associated with improvement in constipation.

**Conclusion:** Anterior active contact locations are associated with improvement in constipation in PD patients after STN DBS. PD patients with greater GCA scores before surgery were more likely to experience urinary deterioration after DBS.

## Introduction

Urinary dysfunction and constipation are two common features of autonomic dysregulation in Parkinson’s disease, occurring in up to 71% and 80% of patients, respectively [1]. Detrusor overactivity is the hallmark of urinary dysfunction in PD [2]. Pelvic floor dyssynergia and colonic motility alteration both contribute to PD constipation [3]. Besides dopamine depletion in the basal ganglia, degeneration of other cortical and subcortical structures may contribute to urinary dysfunction and constipation in PD [1, 4, 5].

Subthalamic nucleus deep brain stimulation (STN DBS), a well-established surgical procedure for PD, significantly improves motor symptoms, but the clinical impact of STN DBS on urinary dysfunction and constipation is variable and unpredictable [1]. Moreover, the neural mechanisms of STN DBS on urinary dysfunction and constipation is undetermined.

The impact of ventromedial and dorsomedial striatum neurostimulation on the visceral contraction and micturition reflex was compared in a murine study: while stimulation of the ventromedial striatum suppressed the micturition reflex, dorsomedial stimulation increased bladder contractions [6]. However, the role of neurostimulation on different STN subregions in humans has not yet been studied.

SPECT studies reveal a correlation between striatum degeneration and bladder dysfunction in PD, particularly in the caudate nucleus [7, 8]. Regional cerebral cortical atrophy also correlates with the severity of urinary dysfunction in PD independent of disease duration [9]. These findings suggest that multivariate surgical and non-surgical components impact the outcome of STN DBS in urinary dysfunction and constipation.

In this study, we investigated the role of active electrode contact location and global cerebral atrophy on the interindividual variability of urinary dysfunction and constipation following STN DBS for PD. We hypothesized that higher global cerebral atrophy score may predict worsened urinary and constipation scores following STN DBS, and stimulation of STN non-motor subregions may improved urinary and constipation scores.

## Method

### Participants and Study Design

Seventy-four patients with a diagnosis of idiopathic PD were included in this retrospective analysis of a prospective database of patients undergoing DBS. All underwent STN DBS at the University of Michigan Surgical Therapies Improving Movement (STIM) program between 2009 and 2019. Written informed consent was obtained from all subjects, and the study was approved by IRBMED, the university institutional review board.

### Surgical Procedure

STN DBS was performed in two stages. For the majority of patients, the first surgery stage involved bilateral implantation of quadripolar DBS electrodes into the STN, with a small number undergoing unilateral implantation. During surgery, microelectrode recording was used to map the STN borders. The tip of the DBS lead was then placed close to the ventral border of the electrophysiological STN. Symptom improvement and side effect thresholds were evaluated intraoperatively by a movement disorders neurologist. During the second stage, an implantable pulse generator was connected to the DBS leads through extension wires. Initial programming of the DBS system was conducted approximately 4-6 weeks after the first stage of surgery. Previous publications have documented our DBS procedure in detail [10].

### Localization of STN Midpoint and Electrode Active Contact

We developed a nuanced method to localize the active electrode contact within STN, considering the interindividual variability of STN shape and size. This method utilizes the STN midpoint coordinate as a reference instead of the midcommissural point [11].

Preoperatively, all patients underwent a 3-T MRI, including a high-resolution coronal imaging protocol that optimized STN visualization [12]. On the day of surgery, patients were fitted with a Leksell stereotactic frame (Elekta Instruments AB, Stockholm, Sweden) and underwent a 1.5-T MRI. The 3-T MRI and 1.5-T MRI were coregistered using a mutual-information algorithm (Analyze 9.0; AnalyzeDirect, Inc, Overland Park, Kansas) and then oriented in Talairach coordinate space along the intercommissural and midsagittal planes. The oral and caudal poles of the STN were selected on a coronal projection of the coregistered MRI. The STN midpoint was defined as halfway between the STN oral and caudal poles [11].

After any brain shift and pneumocephalus resolved at approximately 2-4 weeks following the surgery, a CT scan (GE HD750, 64-slice, kV: 140, mA: 450, 0.5 × 0.5 × 0.6 mm) was obtained to visualize electrode contacts. Coordinates of the active contacts were then determined relative to the MR-visualized STN midpoint. Lateral (X), anterior (Y), and dorsal (Z) directions relative to the STN midpoint were defined as positive.

### Clinical Assessments

Clinical parameters were explored by applying MDS-UPDRS Part I and III before (“off” medication) and 6-12 months after the surgery (“off” medication and “on” stimulation). Urinary problems (item 10) and constipation (item 11) were extracted from Part I. The score range for each item is from 0 (no problems present) to 4 (severe problems present). Demographic information was collected, including age at the follow-up visit, disease duration, gender, and levodopa equivalent dose (LED) at baseline and follow-up visit.

### Measurement of Global Cortical Atrophy

Global Cortical Atrophy (GCA) is a visual rating scale to quantify brain atrophy. Visual rating scales focus on brain regions susceptible to atrophy [13]. GCA evaluates atrophy in 13 regions, including frontal, temporal, and parietooccipital sulcal dilation in each hemisphere and ventricular dilation in the frontal, temporal, and parietooccipital horn of the lateral in each hemisphere and third ventricle [14]. Each region is scored separately from 0 (normal) to 3 (severe), and the final score is the sum of all scores in the 13 regions. GCA was scored for all patients in this analysis, by one clinician for uniformity, using a 3-T MRI FLAIR sequence.

### Statistical Approach

Ordinary Least Square (OLS) model was used to estimate the role of active contact location and GCA score on urinary problems and constipation follow-up score. The forward variable selection method was applied to choose the confounding variables, among LED changes, motor improvement, age, disease duration, gender, and the MDS-UPDRS urinary problems and constipation baseline scores. The cut off value in univariate regression was p<0.25 [15]. Regression analyses were conducted separately for the left and right hemisphere and for urinary problems and constipation, with active contact location predictors separated by axis, resulting in a total of 12 analyses ([urinary problems or constipation] x [right or left hemisphere] x [X, Y, or Z axis]).

To evaluate pre- to post-DBS changes, the score changes in MDS-UPDRS Part I and III were analyzed with repeated measures *t*-tests. Pearson correlation examined the relationship between GCA score with age and disease duration.

### Localization of Maximal-effect Stimulation Loci

Using our previously developed atlas-independent electric field computational model [16, 17], we determined the maximal-effect stimulation loci for constipation and urinary dysfunction and compared these to motor improvement loci. For each condition, patients experiencing an improvement or decline in MDS-UPDRS score were identified. For each condition, a score was generated at each location in space, in a region representing the STN, by using the change in MDS-UPDRS score, the location of the electrode active contact, and the probability of neuronal activation at the location, which utilized factors such as stimulus voltage and distance from the active contact. This weighted score at each location was generated and summed for all patients in a given condition. A simplex algorithm was then used to identify the site associated with the greatest change. Using bootstrapping and repeating this analysis, confidence intervals representing the loci of stimulation leading to maximal change for each condition were generated. In total, four loci were identified that were associated with the greatest improvement and decline for constipation and urinary dysfunction. To assess for statistical significance, the 95% confidence intervals for each condition in each spatial axis were compared, and nonoverlapping confidence intervals (CIs) were determined to be statistically distinct.

## Results

### Participants

Among the 74 patients with idiopathic PD, 72 patients underwent bilateral STN DBS and 2 underwent unilateral left STN DBS. The average patient age was 64.2 ± 7.7 years, with a disease duration of 11.3 ± 4.6 years. The sample population consisted of 22 females. LED decreased by an average of 785.92 ± 540.16 mg post-DBS.

MDS-UPDRS Part III total scores significantly improved following STN DBS (p<0.05) with an average change of 32.74% + 29.34. However, the pre- to post-DBS score changes for urinary problems and constipation were not statistically significant (Table 1).

**Table 1.**

| | Number of Patients | | | | Mean Score ± SD | | P-value |
| --- | --- | --- | --- | --- | --- | --- | --- |
|  | Total | Improved | Deteriorated | Unchanged | Baseline | Follow-up |  |
| Urinary Dysfunction | 74 | 23 | 28 | 23 | $1.08 \pm 0.97$ | $1.18 \pm 1.05$ | 0.49 |
| Constipation | 74 | 25 | 15 | 34 | $1.05 \pm 0.99$ | $0.87 \pm 0.96$ | 0.15 |

### GCA

The GCA score range was from 0 to 31, with an average of 10.81 ± 6.98. Seven patients had no evidence of atrophy (score=0) in all regions, 30 patients had mild atrophy (score=1) in at least one region, 35 patients had moderate atrophy (score=2) in at least one region, and 2 patients had severe atrophy (score=3) in at least one region. GCA score and age were significantly correlated (R=0.47, *p*<0.001), but GCA and disease duration were not (R=0.12, *p*=0.30).

### Active Electrode Contact Locations

On average, the active electrode contact was positioned in the medial (right: -0.15 ± 1.61 and left: -0.42 ± 1.62), posterior (right: -0.95 ± 1.88 and left: -0.42 ± 1.99), and dorsal (right: 1.24 ± 2.04 and left: 1.10 ± 1.90) direction relative to the STN midpoint. The range of distance to STN midpoint was from -6 to 3 mm on X-axis, -5.5 to 4 mm on Y-axis, and -3 to 8 mm on Z-axis.

### Association between Urinary Problems and Active Electrode Contact and GCA

GCA score was a significant predictor in the same direction for urinary dysfunction follow-up scores (*B* = 0.34, *p* = 0.01), while active contact locations were not significant predictors (*p*>0.05 in all six models). Among the confounding variables, age and motor improvement percent change were included in the main model (Table 2). In the final model, age and GCA score independently explained 38% of urinary follow-up score variance (R^2^ = 0.38. *p<*0.001).

**Table 2.**

| **Table 2** |  |  |  |  |
| --- | --- | --- | --- | --- |
| *Forward Variable Selection* |  | *Main Model (N=74)* |  |  |
| Confounding Variable | *p*-value | Selected Variables | *B* (standardized coefficient) | *p*-value |
| Urinary baseline score | 0.31 | Age | 0.34 | 0.01 |
| Motor improvement percent change | 0.07 | GCA | 0.40 | 0.00 |
| LED change | 0.45 | Motor improvement percent change | 0.05 | 0.64 |
| Age | 0.00 |  |  |  |
| Disease duration | 0.26 |  |  |  |
| Gender | 0.42 |  |  |  |

### Association between Constipation and Active Contact Locations and GCA

A more anterior active contact location within the STN in both right and left hemispheres was a significant predictor for the improvement of constipation follow-up score. GCA score did not predict the constipation follow-up score (Table 3). Among the confounding variables, disease duration, constipation score at baseline, and motor improvement percent change were included in the main model. The final model explained 33% of constipation follow-up scores variance (R^2^ = 0.33, *p<*0.001). Table 3 presents the standardized coefficient

**Table 3.**

| *Forward Variable Selection* |  | *Main Model Left Hemisphere (N=74)* |  |  | *Main Model Right Hemisphere (N=72)* |  |  |
| --- | --- | --- | --- | --- | --- | --- | --- |
| Confounding Variables | *P-value* | Selected Variables | *B* | *P-value* | Selected Variables | *B* | *P-value* |
| Constipation baseline score | 0.00 | Active contact in Y-axis | -0.28 | 0.00 | Active contact in Y-axis | -0.23 | 0.00 |
| Motor improvement percent change | 0.04 | Constipation baseline score | 0.34 | 0.00 | Constipation baseline score | 0.37 | 0.00 |
| LED change | 0.88 | Motor improvement percent change | -0.13 | 0.20 | Motor improvement percent change | -0.12 | 0.23 |
| Age | 0.43 | Disease Duration | 0.24 | 0.03 | Disease Duration | 0.23 | 0.03 |
| Disease duration | 0.00 | GCA | 0.09 | 0.36 | GCA | 0.05 | 0.57 |
| Gender | 0.84 |  |  |  |  |  |  |

### Maximal-effect Stimulation Loci

We found the maximal-effect stimulation loci for constipation improvement and deterioration were significantly distinct without overlap between CIs on the Y-axis (Table 4). Figure 1 depicts the maximal-effect loci for constipation within an exemplar traced STN. For urinary dysfunction, there was an overlap between CI in all axes (Figure 2).

**Table 4.**

|  | X-axis | Y-axis | Z-axis |
| --- | --- | --- | --- |
| Constipation Improvement | -0.25 [-0.85, 0.25] | **0.00 [-0.42, 0.49]** | 1.14 [0.58, 2.08] |
| Constipation Deterioration | -0.52 [-1.35, 0.07] | **-2.03 [-2.82, -.71]** | 0.68 [0.00, 1.55] |
| Urinary symptoms Improvement | -0.29 [-0.91, 0.20] | -0.34 [-1.04, 0.35] | 1.75 [0.90, 2.81] |
| Urinary symptoms Deterioration | -.41 [-.84, 0.00] | -1.14 [-1.85, -0.38] | 1.07 [0.43, 1.89] |
| Motor Improvement | -0.22 [-0.5, 0.04] | -0.70 [-1.10, -0.31] | 0.98 [0.63, 1.34] |

Anterior, lateral, and dorsal are positive. All values are in millimeters.

**Figure 1.**
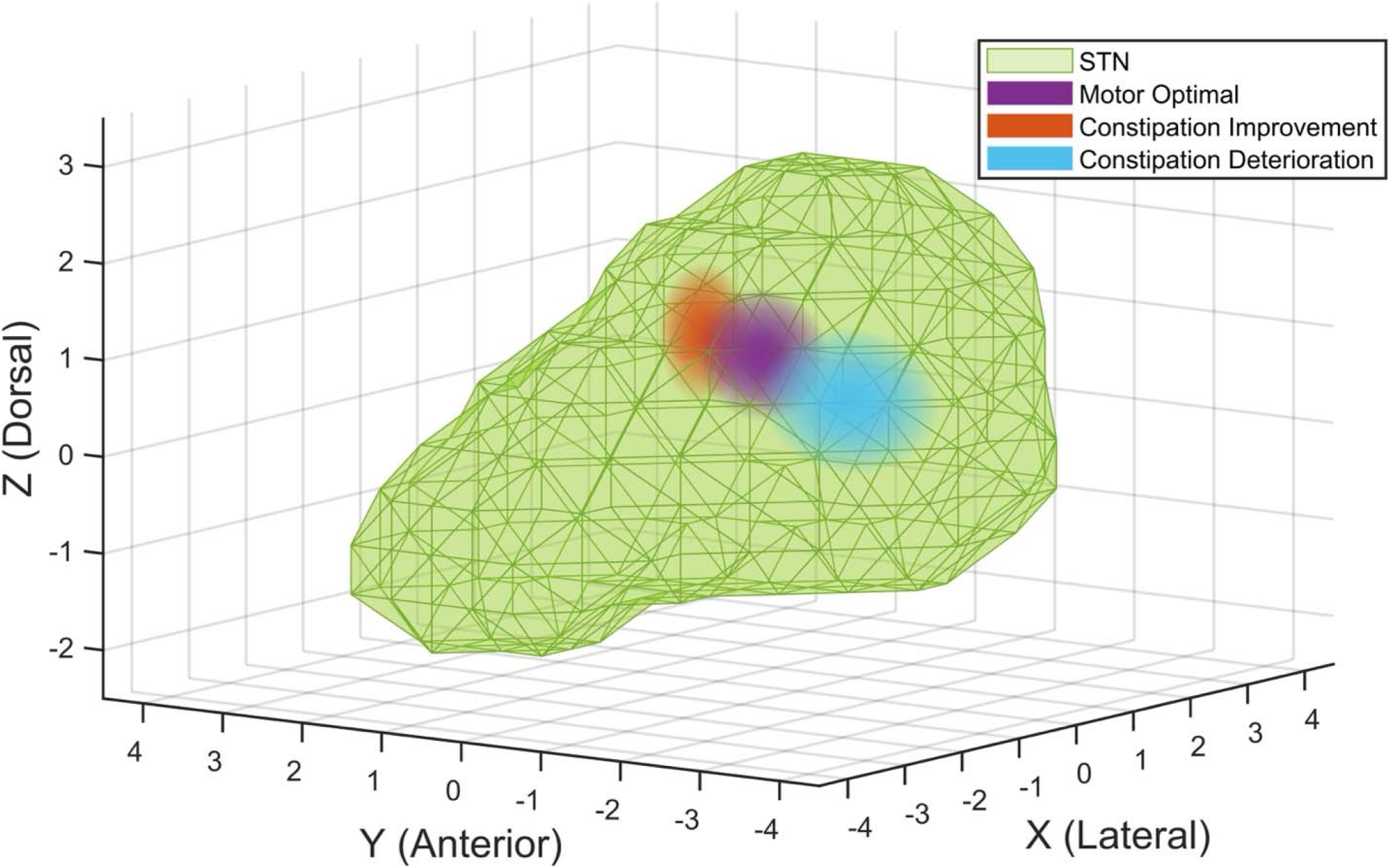
Loci of motor and constipation effects for STN DBS stimulation. Ninety-five percent confidence intervals for motor improvement (violet), constipation improvement (red), and constipation deterioration (blue). Regions for constipation improvement and deterioration improvements are statistically distinct.

**Figure 2.**
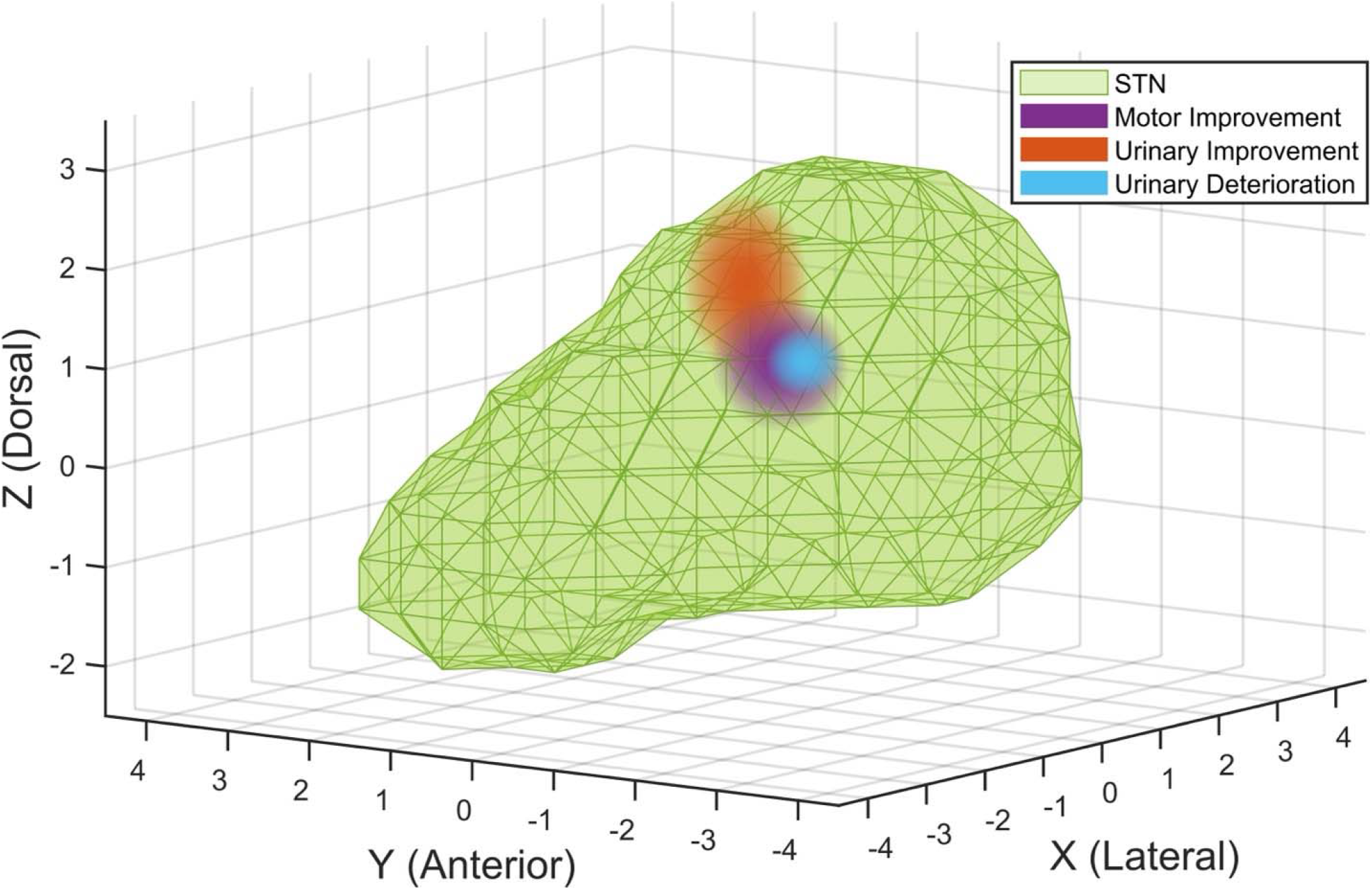
Loci of motor and urinary effects for STN DBS stimulation. Ninety-five percent confidence intervals for motor improvement (violet), urinary improvement (red), and urinary deterioration (blue). Regions for urinary improvement and deterioration improvements are not statistically distinct.

## Discussion

When the cohort was analyzed as a whole, neither urinary dysfunction nor constipation scores changed on average following STN DBS. However, there was significant interindividual variability with some individuals reporting improvement in urinary symptoms and/or constipation while others reported a deterioration. We found that active contact stimulation anterior to the STN midpoint in each hemisphere predicted improvement in constipation at follow-up after adjusting for disease duration, motor improvement, and constipation baseline score. The locus of maximal-effect stimulation for constipation improvement vs deterioration in the Y-axis was distinct and the locus of motor improvement was placed between them (Figure 1). Additionally, a greater baseline cerebral atrophy, as evidenced by a higher GCA score, was associated with increased urinary problems after adjustment for age.

The beneficial effect of STN DBS on urinary symptoms and constipation has been shown individually [18, 19]. However, there are cases with significant worsening of symptoms following STN DBS surgery [20, 21]. The varied results suggest a complex interplay between DBS and non-surgical parameters, including disease duration, age, medication, and cerebral atrophy [1].

Nigrostriatal dopaminergic and non-dopaminergic degeneration is associated with bladder overactivity and the severity of urinary symptoms in PD. Wing and colleagues evaluated the degeneration of nigrostriatal structures using SPECT imaging and showed a correlation between the severity of bladder dysfunction and caudate nucleus degeneration. [7]. In their study, however, the loss of striatal neurons did not predict the effects of dopaminergic medication on bladder control. They therefore postulated that structures outside of basal ganglia might be involved [7]. In a recent study, corticometric and volumetric analyses of various cortical regions illustrated a correlation between the severity of urinary dysfunction in PD and decreased cortical thickness and volume, especially in the precuneus cortex [9].

Our study is the first to date to evaluate the impact of global cerebral atrophy on the urinary symptoms following STN DBS. GCA scale is a visual rating scale applied to patients with dementia that improved the sensitivity and reliability of radiological image interpretation [13]. The determination of atrophy in different brain regions increases the diagnostic value. [13], and the GCA scale has been validated in several volumetric studies [22]. Cerebral atrophy, large lateral ventricles, and structural pathology have been studied as preoperative imaging factors that may influence the outcome of deep brain stimulation in PD [23]. However, the evidence was insufficient; some centers avoid surgery in patients with extensive cerebral atrophy [23], [24]. This study demonstrates that the magnitude of global brain atrophy impacts urinary symptoms following DBS.

The pathophysiology of constipation is complex, and the dorsal motor nucleus of the vagus nerve has been implicated in PD constipation [1]. Braak and colleagues posited that alpha-synuclein was initially deposited in the dorsal motor nucleus of the vagus nerve and anterior olfactory nucleus [25], which would be consistent with the clinical observation that constipation can predate PD diagnosis [26].

Pagano and colleagues, applying MDS-UPDRS Part I and II, suggested constipation as a pre-motor symptom of PD with a non-dopaminergic pathway [4]. In a murine study, Derry and colleagues showed that unilateral STN DBS significantly increased colonic motility [3] and by applying immunochemistry techniques, demonstrated that this occurred by activation of the dorsal motor nucleus of the vagus nerve [3]. They hypothesized that STN DBS may activate cortical limbic areas that modulate colonic behavior and project to locus coeruleus and efferent spinal pathways [3] [27]. The anterior STN has both anatomical and functional connectivity with limbic structures [28], which would be consistent with our finding that stimulation of the anterior subregion of STN improves constipation follow-up score.

In the recent large cohort study, Camacho and his colleagues, applying MDS-UPDRS Part I and II, showed the association between constipation severity and progression to dementia. They postulated that constipation might have a prognostic role in PD and could be a target for disease modification [29]. Poewe and his colleagues showed the relationship between autonomic dysfunction with dementia and cognitive dysfunction in PD. They hypothesized that Lewy body pathology in limbic areas that cause cognitive dysfunction [30, 31] may also impact autonomic dysfunction [32]. In light of our findings, further investigation of neurostimulation of limbic areas within the anterior STN subregion and their connections and its effects on constipation and cognitive dysfunction is warranted.

## Limitations

This study has several limitations. We exclusively used MDS-UPDRS Part I questionnaires items 10 and 11 to evaluate urinary symptoms and constipation. While these items have been validated with autonomic scales, [33], [34] evaluating urinary symptoms and constipation with more specific detailed questionnaires or urodynamic studies could provide more detailed information. We did not incorporate electrical DBS parameters into the regression analysis and did not calculate volume of tissue activation which has been helpful in determining the optimal site for motor improvement [35]. Finally, we used GCA, a visual scale instead of using automated volumetric MRI analysis, which is compared to age-matched controls.

## Conclusion

STN DBS heterogeneously impacts urinary dysfunction and constipation, varying from deterioration to improvement. Anterior STN stimulation improves constipation scores, while the locus of stimulation is not associated with changes in urinary outcomes. Non-surgical factors, including age and cerebral atrophy, worsened urinary symptoms post-DBS. These factors may be considered when assessing STN DBS candidacy in PD.

## Data Availability

All data produced in the present study are available upon reasonable request to the authors and IRB approval

